# Trends in psychological distress in Great Britain, 1991-2019: evidence from three representative surveys

**DOI:** 10.1101/2022.08.08.22278544

**Authors:** Anwen Zhang, Thierry Gagne, David Walsh, Alberto Ciancio, Eugenio Proto, Gerry McCartney

## Abstract

**Background:** Previously improving UK mortality trends stalled around 2012 due to economic policy changes. This paper examines whether trends in psychological distress across three population surveys show similar trends.

**Methods:** We report the percentages reporting psychological distress (4+ in the 12-item General Health Questionnaire (GHQ-12)) from Understanding Society (Great Britain, 1991-2019), Scottish Health Survey (SHeS, 1995-2019) and Health Survey for England (HSE, 2003-2018) for the population overall, and stratified by sex, age and area deprivation. Summary inequality indices were calculated and segmented regressions fitted to identify turning points after 2010.

**Results:** Psychological distress was higher in Understanding Society than in the SHeS or HSE. There was a slight improvement between 1992 and 2015 in Understanding Society (with prevalence declining from 20.6% to 18.6%) with some fluctuations. After 2015 there is some evidence of an overall deterioration in psychological distress across surveys. Prevalence worsened notably among those aged 16-34 years after 2010 (all three surveys), and aged 35-64 years in Understanding Society and SHeS after 2015. In contrast, the prevalence declined in those aged 65+ years in Understanding Society after around 2008, with less clear trends in the other surveys. The prevalence was around twice as high in the most deprived compared to the least deprived areas, and higher in women, with trends by deprivation and sex similar to the populations overall.

**Conclusion:** Psychological distress worsened amongst working-age adults after around 2015 across British population surveys, mirroring the mortality trends. This indicates a widespread health crisis that pre-dates the COVID-19 pandemic.

## Introduction

From the mid-20^th^ Century onwards, average life expectancy in the UK increased steadily. ^1^ However, these average trends stalled after around 2012 across all UK nations, ^2 3^ and mortality rates increased for those living in the most deprived areas. ^4 5^ There is evidence that healthy life expectancy, a measure that combines self-rated health with mortality, has also been declining in Scotland over a similar time period. ^6^ However, it is less clear whether trends in mental health and wellbeing have changed in tandem with mortality trends.

Unlike all-cause mortality, ^7^ it is known that mental health is likely to worsen during periods of recession. ^8^ However, mortality trends, and mental health trends, may be impacted more by the economic policy responses to recession rather than recessions themselves. ^7 9^ Indeed, the austerity policies imposed by a range of countries following the ‘Great Recession’ (c.2008-2010) has been demonstrated as the likely explanation for the stalled mortality trends. ^9 10^

Routine surveillance data from the Scottish Health Survey (SHeS) and Health Survey for England (HSE) are available to examine annual trends across the UK using the General Health Questionnaire (GHQ-12), a longstanding screening tool for identifying non-psychotic and minor psychiatric disorders in the general population. The Understanding Society panel dataset also provides a means to monitor mental health trends. Although there have been analyses of particular population sub-groups, ^11 12 13 14 15 16^ there has not yet been any comprehensive analyses of trends in psychological distress for the population overall, sex, age and deprivation strata, nor has there been any comparison across concurrent survey datasets for these populations.

This paper explores whether trends in mental health changed in parallel to the overall life expectancy trends using population-representative data from the Understanding Society, from 1991 to 2019; Health Survey for England (HSE) from 2003 to 2018 and Scottish Health Survey (SHeS) from 1995 to 2019, using the prevalence of psychological distress based on the GHQ as the outcome of interest.

## Methods

### Data sources

GHQ-12 is a 12-question validated survey measure used to estimate the prevalence of psychological distress in populations. ^17^ GHQ-12 prevalence represents the share of the population with a score of 4+, on a scale between 0 and 12, with higher values indicating greater psychological distress. The GHQ questionnaire is presented in Supplement 1.

We analyse data for people aged 16+ years from:

1. Understanding Society, from 1991 to 2019 (called the British Household Panel Survey (BHPS) from 1991 to 2008, and the UK Household Longitudinal Survey until 2019). The data are weighted to represent Great Britain (GB).
2. Health Survey for England (HSE) from 2003 to 2018 (with gaps for some years). The data are weighted to represent England.
3. Scottish Health Survey (SHeS) from 1995 to 2019 (with gaps for some years). The data are weighted to represent Scotland.

### Analytical approach

Simple descriptive figures for the total weighted populations, and then stratified by sex, age and area deprivation fifth, were produced. The summary inequality indices, the Slope Index of Inequality (SII) and Relative Index of Inequality (RII) were produced assuming that each deprivation fifth represented exactly 20% of the population. Using the approach detailed by Pamuk, ^18^ we then produced linear regressions for each year across the deprivation categories to calculate the SII, and divided that by the population mean to calculate the RII. Finally, we fitted segmented regression models to the data trends in each survey, age group and deprivation fifth, limiting the number of splines (or turning points) to one, and to the period after 2010.

## Results

### Overall population trends

Figure 1 shows trends in the percentage of the total adult populations of GB, Scotland and England scoring 4+ on the GHQ-12 between 1991 and 2019. The time series for GB from the Understanding Society dataset is the longest and displays a gradual improvement (i.e. a decline in the percentage reporting psychological distress), albeit with substantial year-on-year variation, between 1992 and 2015. However, after 2015 the percentage of people reporting psychological distress steadily increases to 2019. The Scottish Health Survey (SHeS) shows a similar trend, but with substantially lower percentages until 2018. Here, the percentage is relatively stable until around 2016, with a subsequent increase peaking in 2018. The Health Survey for England (HSE) figures are similar to those from SHeS, and lower than Understanding Society. Here there is some evidence of an increasing trend over time, but with peaks in 2009 and 2016. The trends are very similar when the data are stratified by sex, with similar increases across the surveys after 2015. However, the percentage of the population reporting psychological distress is consistently higher for women than men (Figures S2.1 and S2.2).

**Figure 1.**
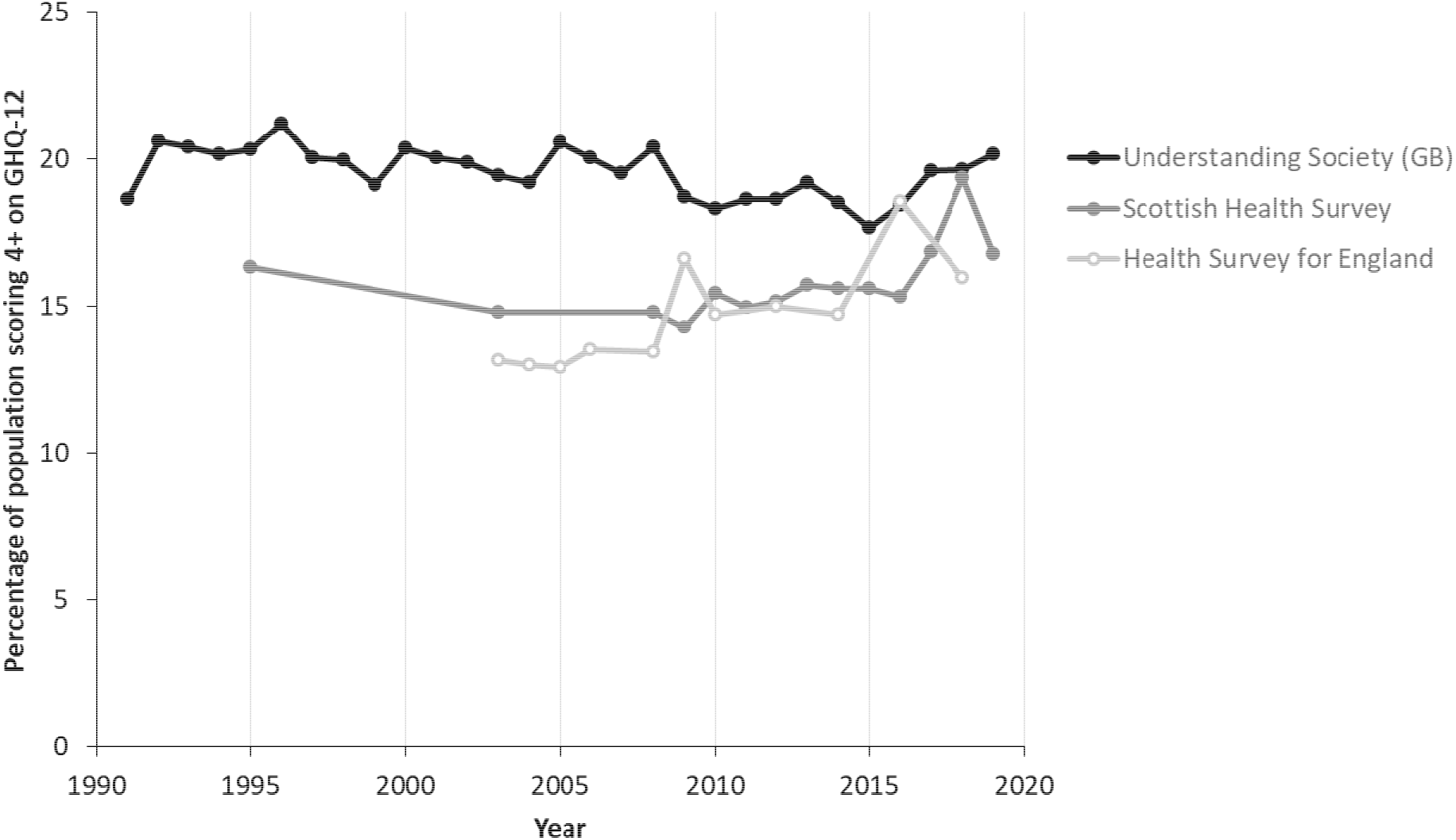
Trends in psychological distress in GB, Scotland and England, 1991-2019 (Sources: Understanding Society, Scottish Health Survey, Health Survey for England)

### Age-stratified trends

When the trends are stratified by age, there is a marked divergence which is obscured in the average population data (Figure 2), although the smaller sample sizes mean that the trends have more year-on-year variation than for the total population. The increase in psychological distress after 2015 is only seen amongst people aged 16-34 and 35-64 years. For those aged 65+ years, there is a decline in psychological distress after 2008 in Understanding Society, but with less stable trends in SHeS and HSE (indeed, HSE shows a marked peak in 2009 for this age group). It is also notable that the increase in psychological distress is evident earlier for those aged 16-34 years compared to those aged 35-64 years, starting around 2010.

**Figure 2.**
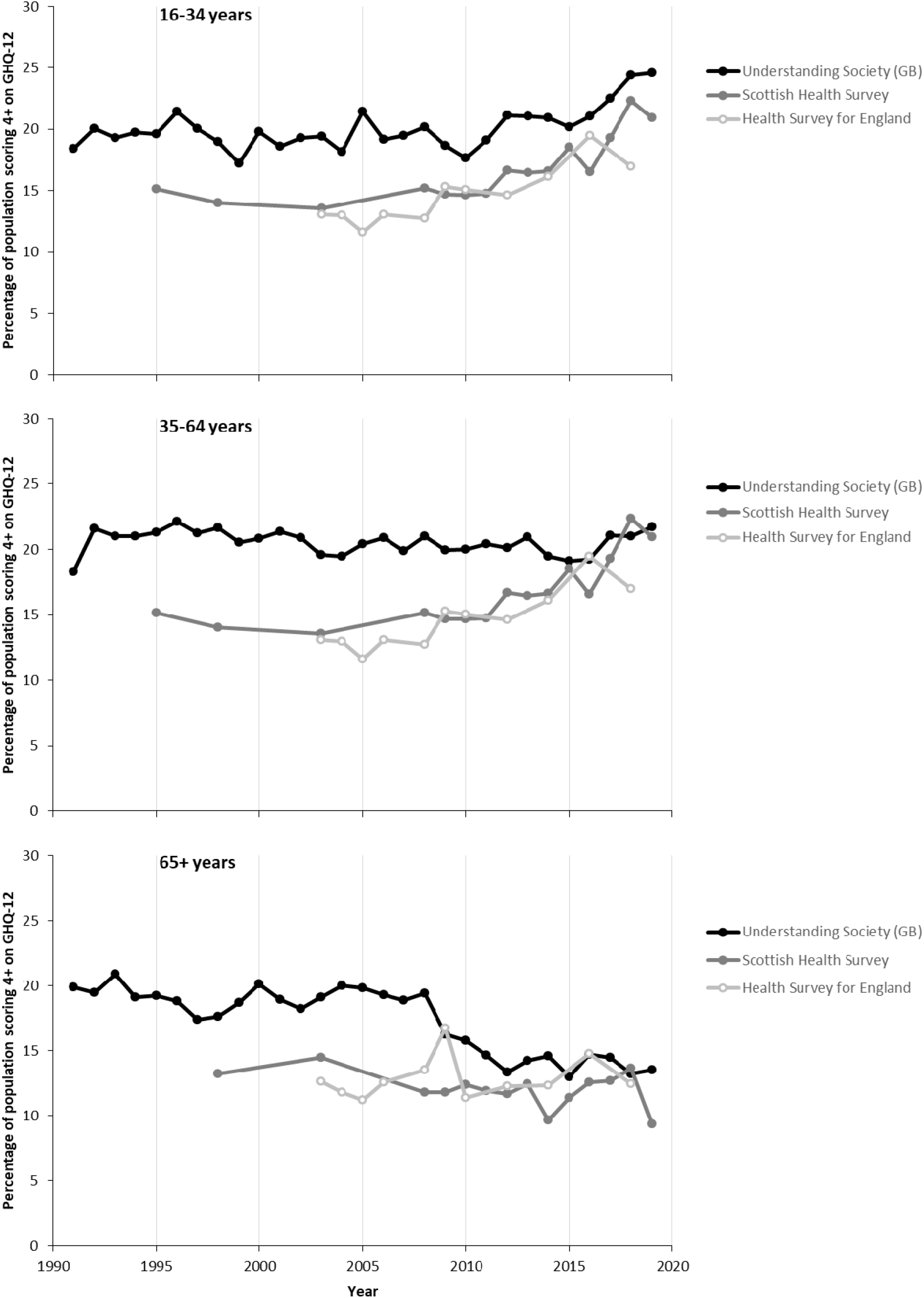
Trends in psychological distress in GB, Scotland and England, stratified by age, 1991-2019 (Sources: Understanding Society, Scottish Health Survey, Health Survey for England)

### Deprivation-stratified trends

When the data are stratified by area deprivation, the inequalities in psychological distress are exposed (Figure 3). The trends in the summary measures of absolute (Slope Index of Inequality, SII) and relative (Relative Index of Inequality, RII) inequality across the populations are provided in Figures S2.3-S2.5. Across GB, Scotland and England, and for the entire time series, there are large inequalities in the prevalence of psychological distress, with a stepwise gradient across the ranked population. The trends for the SII and RII are very similar across all datasets as there is no strong secular trend in the population means (which would create a divergence between the absolute and relative measures). In the SHeS and HSE, the percentage of the population reporting psychological distress in the most deprived areas is around double that of the least deprived areas. The stepwise gradient across the population becomes clearer in the Understanding Society dataset only after 2009, and is associated with a notable increase in inequalities (Figure S2.3) that looks likely to be associated with the changes in the panel make-up at that time. Prior to 2009 there are some marked year-on-year fluctuations in inequalities in the Understanding Society dataset, and evidence of reduced inequalities after 2013. Within the SHeS and HSE datasets the trends in inequalities after 2010 are very unstable with marked fluctuations between years on both the SII and RII reflecting a lack of parallel trends across deprivation fifths. It is notable that the rise in the percentage reporting psychological distress in Scotland is much more marked in the most deprived 40% of areas.

**Figure 3.**
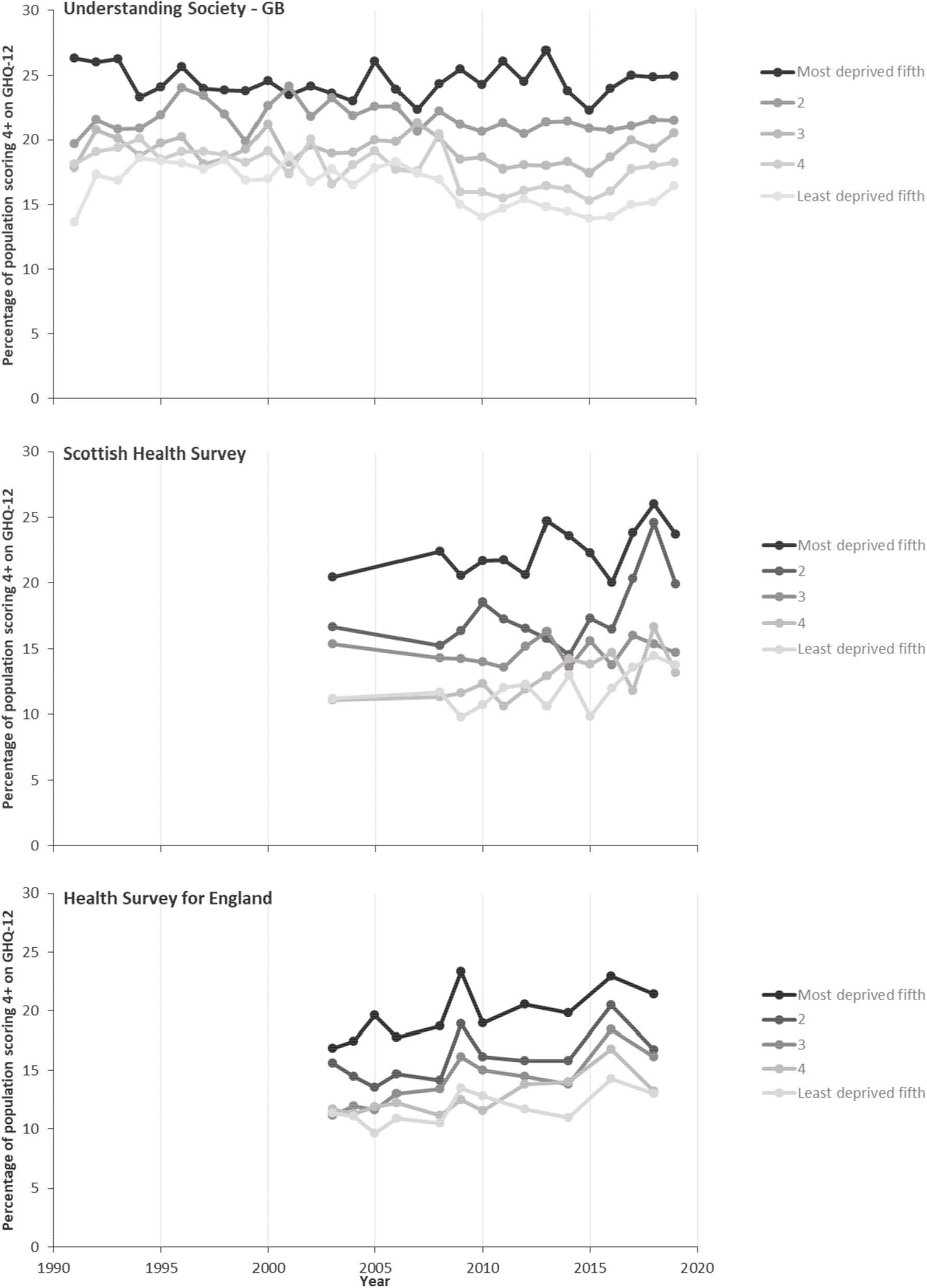
Trends in psychological distress in GB, Scotland and England, stratified by area deprivation, 1991-2019 (Sources: Understanding Society, Scottish Health Survey, Health Survey for England)

### Intersectional stratifications

When the data are stratified concurrently across combinations of age, sex and deprivation, the numbers in each group are much smaller, and as a result the trends become less stable and clear. Figures S2.6 and S2.7 show the trends stratified by deprivation and sex, and indicate clearer and wider inequalities by deprivation for men than women. The deprivation inequalities are widest amongst those aged 35-64 years compared to younger or older adults (Figures S2.8-S2.10).

### Turning points in the trends

When formal segmented regression analyses are fitted to the data to identify the turning points in the trends, there is a change from improving trends to a marked worsening in the trends for Scotland (SHeS) after 2011, and across GB after 2016 (Understanding Society) that are obvious. The trend for England worsens throughout the time series with only a minor change in trend (towards a less rapid worsening between 2016-18) (Figure S2.11). The changes in trends and their timing are similar when the data are stratified by sex (Figure S2.12). However, when the data are stratified by age the differences noted in the descriptive analyses are obvious. For people aged 16-34 years and 35-64 years psychological distress worsens in the latest time period, and except for the HSE data for 35-64 year olds, are getting worse more quickly. The change in trends for 16-34 year olds occurs earlier (c.2010, across surveys) than for people aged 35-64 year old, and especially in Understanding Society. In contrast, with the exception of SHeS, there is a more rapid improvement in those aged 65+ years in the most recent period (Figure 2.13). Finally, when the data are stratified by deprivation fifth there is some variance in the timing of the changes in trends (Figure S2.14).

## Discussion

### Main results

The percentage of the overall population scoring 4+ on the GHQ-12 over time is higher in Understanding Society (for GB) than in the SHeS (for Scotland) or HSE (for England). The long time series for GB improved between 1991 and around 2015, albeit with some fluctuations. After 2015 there is some evidence of a deterioration in mental health across surveys, but the data for Scotland and England has substantial year-to-year variation. However, there are much clearer trends when the data are stratified by age, with consistent increases in psychological distress across surveys for those aged 16-34 years after 2010, and increases for those aged 35-64 years in Understanding Society and SHeS after 2015. In contrast, psychological distress declines in those aged 65+ years in Understanding Society after around 2008, and shows no clear pattern in the other surveys. Throughout the time series there are substantial inequalities in psychological distress with around double the prevalence in the most deprived areas compared to the least deprived areas, and a higher prevalence amongst females compared to males.

### Strengths and limitations

Although life expectancy is a critical measure of the health of populations, it is also a very narrow one. Taken with the existing work on mortality and healthy life expectancy,^2 3 4 5 6 9 19 20 21^ this paper provides a more holistic picture of health and health inequality trends and the extent to which the stalling of life expectancy is reflected in other measures.^22^ The data used here are among the best available for understanding the experience of mental health. They are drawn from government funded surveys which aim to be representative of the population, and use validated and comparable measures of the prevalence of psychological distress. ^17^ By triangulating data across three surveys there is greater certainty in the trends, and the difficulties associated with the periodic changes in sampling are reduced. Using survey data avoids the problems associated with changes in clinical practice, or the acceptability of treatments, as would be the case if prescribing for mental health conditions was used as an outcome. Similarly, the outcome is not impacted by health service capacity or focused on only the most severe mental health outcomes as would be the case with admission to hospital or suicide mortality. By stratifying the data we identified divergent trends, particularly by age, which were otherwise obscured.

We limited the Understanding Society dataset to Great Britain to avoid the step change associated with the inclusion of Northern Ireland part way through the time series. However, there was a change in the sample for Understanding Society after 2009 which may have created a risk of a step change. Although the data used here are the best available for GB, Scotland and England, they are subject to limitations. In particular, non-response bias is a growing problem for the SHeS and HSE (and made the data in 2020 and 2021 virtually unusable), and is likely to have created an ever-increasing healthy-responder bias. ^23^ It would be expected that this would bias the sample and the trends towards improving health outcomes over time, and so the increased prevalence of psychological distress might be a substantial under-estimate of the extent of the problematic trend. Some of the secular trends may also have been due to changes in the age structure of the populations over time, particularly given the different trends observed when the data were age stratified.

### How this fits with the existing literature

The stalling in life expectancy trends after around 2012 across the UK, and many other high income countries, has been clearly described.^2 3 4 5 19 20^ This has been due to changes in mortality trends across almost all age groups and causes of death. ^20 21^ More recently a deterioration in healthy life expectancy has also been described, with contributions from both self-rated health and mortality. ^6^ Psychological distress arguably has a non-linear relationship with age, being worst in middle-age. Despite this, within the Understanding Society dataset, the temporal trends have been found to dominate the impacts of cohort or age effects. ^24^ We find here that there are markedly divergent trends by age group across surveys, with much clearer increases in prevalence for younger adults than older adults. As noted previously, recessions may narrow socioeconomic and gender inequalities in mental health, but austerity exacerbates them. ^25^

The worsening in psychological distress amongst young adults in GB described in our study is consistent with other analyses using these datasets, and with data from primary care records. In Scotland, there was an increase in reporting of symptoms of anxiety, depression, and the reporting of suicide attempts and having ever self-harmed, between 2012-13 and 2018-19. ^26^ There does not seem to have been any trend analyses published by the HSE team based on GHQ-12 for all adults after 2010. ^27^ Other researchers have used the HSE and found that, between 1991 and 2010, mental health (measured with GHQ-12) in English working age adults (25-64 years) slightly improved on average, especially between 1997 and 2003 for women,^11^ but with persistently worse mental health for women and for people with lower incomes, less education and in lower socioeconomic positions.^28^ Similarly, amongst studies focused on children and young people in GB, there was evidence of increased prevalence of long-standing mental health conditions between 1995 and 2014, and in psychological distress between 2011 and 2014, ^13^ and between 2009/2010 and 2018/19, particularly for those aged 16-18 years and of white ethnicity. ^14^ The increase in psychological distress amongst young adults (aged 16-34 years) in the British Household Panel Survey/Understanding Society has been reported previously. ^15^ Finally, there is also evidence of a rise in anxiety and depression diagnoses and symptoms amongst people aged 18-24 years, and especially women, from UK primary care records between 2014 and 2018. ^16 29^

Relevant to explaining these trends, a systematic review of observational studies showed that policies which reduced the eligibility and generosity of social security led to a worsening in mental health and increased inequalities, whilst increased eligibility and generosity improved mental health. ^16^ For example, the change to Universal Credit in the UK, which reduced eligibility and the real value of social security, led to a 7% increase in psychological distress after adjusting for confounders. ^30^ Indeed, there is evidence that tightening of the sanctions on unemployment benefits in 2012, which increased the length of time for which people were in receipt of no benefit, led to increases in anxiety and depression. ^31^ There is also now high quality systematic review evidence that changes in income, and especially movement into or out of poverty, have a profound impact on mental health and wellbeing. ^32^ In Scotland, antidepressant use increased over the period 2009 to 2015, and especially in those areas impacted most by unemployment and reductions in the real value of social security benefits. ^33^ More recently, a wide range of mental health measures are evidenced to have worsened during the COVID-19 pandemic for some groups, ^34 35 36 37^ with mixed results for the short-term impacts of the pandemic from studies with less robust sampling methods. ^38 39^

### Implications

The stalling of life expectancy trends and mortality inequalities are amongst the biggest public health challenges we face, ^40^ with very substantial impacts on the Years of Life Lost. ^41^ The stalled life expectancy trends are likely to be largely due to the austerity policies introduced from 2010 onwards, and the associated cuts to local government funding and social security. ^9^ There is now a substantial body of evidence that the increased conditionality and reduced real terms value of social security benefits has had marked deleterious impacts on mental health, and this is likely to be a substantial contributor to the overall population mental health trends exposed here. Governments should act to protect population health and reduce health inequalities by addressing the economic causes of these trends as well as the wider range of social determinants of health, the recommendations for which have been published in detail elsewhere. ^9 40 42 43 44^

## Conclusion

The prevalence of psychological distress increased after 2010 in young adults, and after 2015 for working-age adults across GB. Inequalities in psychological distress are substantial by area deprivation, with around double the prevalence in the most deprived areas compared to the least deprived areas. Psychological distress is higher in females than in males. The trends in psychological distress are similar to those for overall population life expectancy and healthy life expectancy, indicating a population health challenge that predates the COVID-19 pandemic and which demands a radical and coherent governmental response.

## Data Availability

The Understanding Society, Scottish Health Survey, and Health Survey for England datasets used here are available online on the UK Data Service platform.

https://ukdataservice.ac.uk/

## Contributions

EP and GM conceived the study and designed it with input from AZ, TG, DW and AC. AZ, TG and DW obtained and cleaned the data and performed the descriptive analyses. AZ performed the segmented regression analyses. GM drafted the manuscript. All authors provided critical input to the final draft.

