## Supplement 1 for "Trends in psychological distress in Great Britain, 1991-2019: evidence from three representative surveys"

**Supplement 1: The General Health Questionnaire (GHQ) 12**

The next questions are about how you have been feeling over the last few weeks.

- Have you recently been able to concentrate on whatever you're doing?

1. Better than usual

2. Same as usual

3. Less than usual

4. Much less than usual

- Have you recently lost much sleep over worry?

1. Not at all

2. No more than usual

3. Rather more than usual

4. Much more than usual

- Have you recently felt that you were playing a useful part in things?

1. More so than usual

2. Same as usual

3. Less so than usual

4. Much less than usual

- Have you recently felt capable of making decisions about things?

1. More so than usual

2. Same as usual

3. Less so than usual

4. Much less capable

- Have you recently felt constantly under strain?

1. Not at all

2. No more than usual

3. Rather more than usual

4. Much more than usual

- Have you recently felt you couldn't overcome your difficulties?

1. Not at all

2. No more than usual

3. Rather more than usual

4. Much more than usual

- Have you recently been able to enjoy your normal day-to-day activities?

1. More so than usual

2. Same as usual

3. Less so than usual

4. Much less than usual

- Have you recently been able to face up to problems?

1. More so than usual

2. Same as usual

3. Less able than usual

4. Much less able

- Have you recently been feeling unhappy or depressed?

1. Not at all

2. No more than usual

3. Rather more than usual

4. Much more than usual

- Have you recently been losing confidence in yourself?

1. Not at all

2. No more than usual

3. Rather more than usual

4. Much more than usual

- Have you recently been thinking of yourself as a worthless person?

1. Not at all

2. No more than usual

3. Rather more than usual

4. Much more than usual

- Have you recently been feeling reasonably happy, all things considered?

1. More so than usual

2. About the same as usual

3. Less so than usual

4. Much less than usual
