## Supplement 2 for "Trends in psychological distress in Great Britain, 1991-2019: evidence from three representative surveys"

**Figure S2.1 – Female trends in psychological distress in GB, Scotland and England, 1991-2019 (Sources: Understanding Society, Scottish Health Survey, Health Survey for England)**

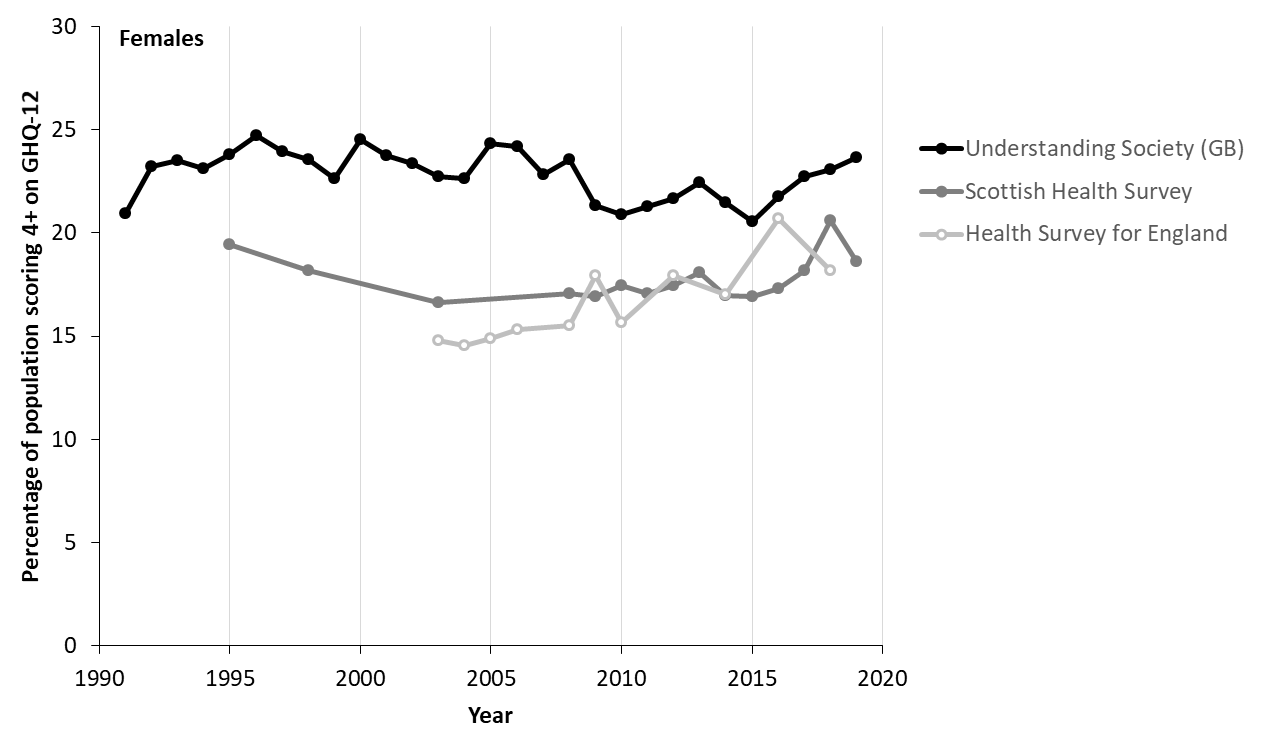

**Figure S2.2 – Male trends in psychological distress in GB, Scotland and England, 1991-2019 (Sources: Understanding Society, Scottish Health Survey, Health Survey for England)**

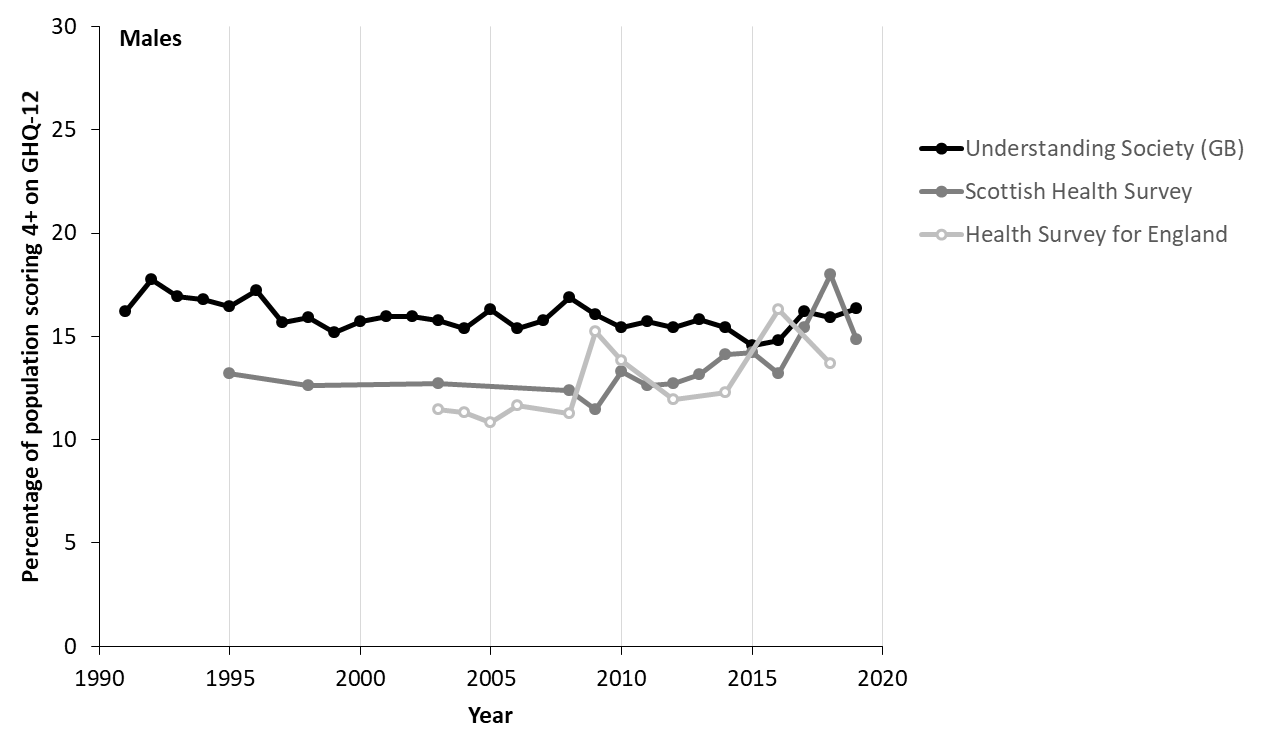

**Figure S2.3 – Trends in area deprivation-based inequalities in psychological distress in GB, measured using the Slope Index of Inequality (SII) and Relative Index of Inequality (RII), 1991-2019 (Source: Understanding Society)**

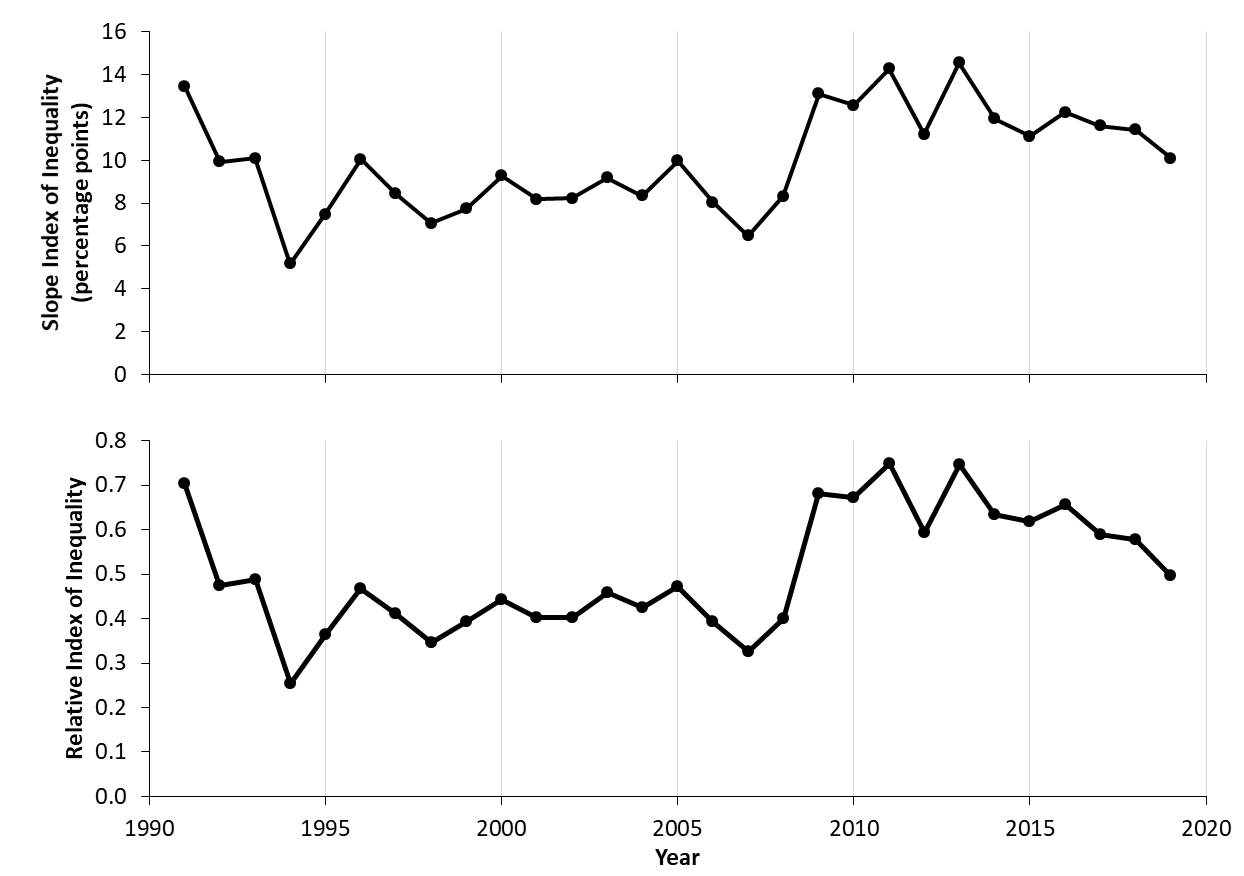

**Figure S2.4 – Trends in area deprivation-based inequalities in psychological distress in Scotland, measured using the Slope Index of Inequality (SII) and Relative Index of Inequality (RII), 1991-2019 (Source: Scottish Health Survey)**

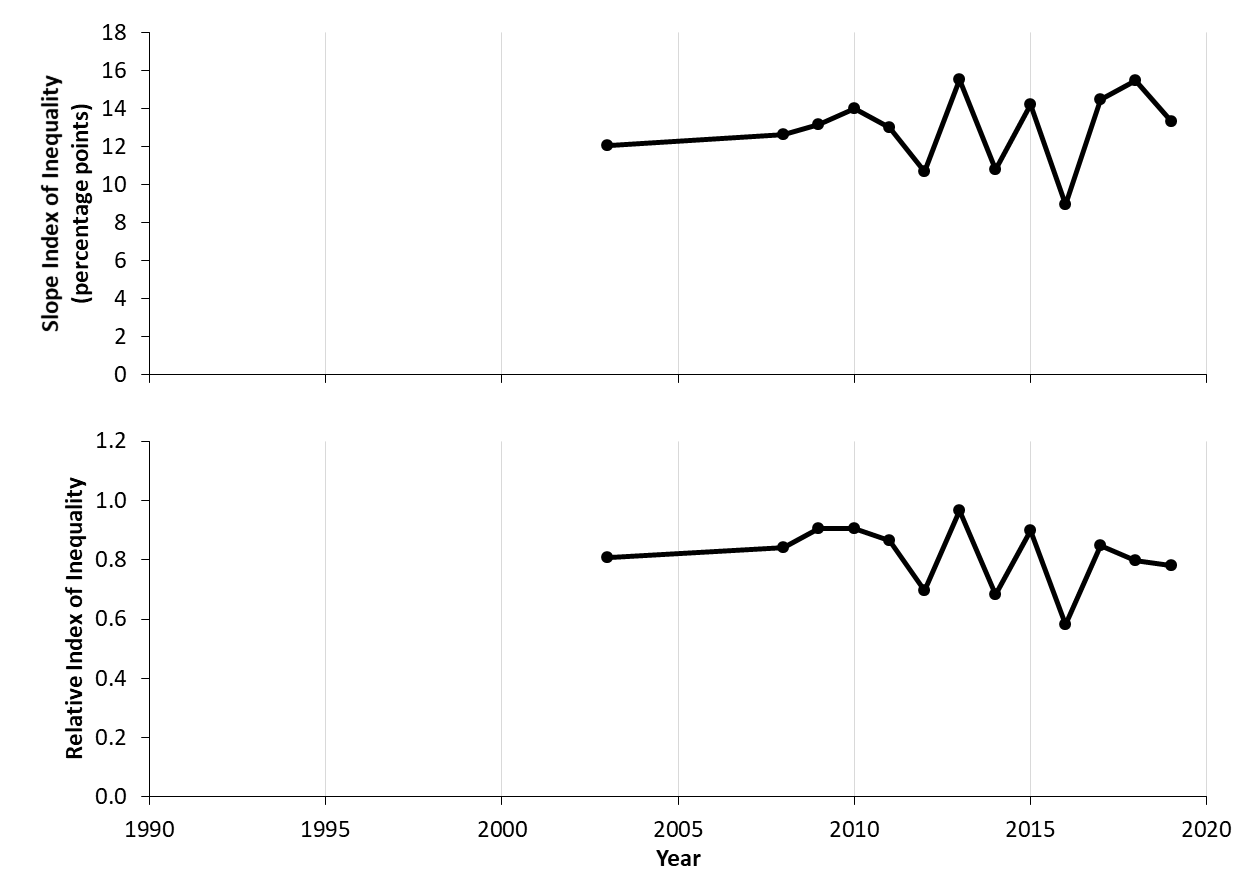

**Figure S2.5 – Trends in area deprivation-based inequalities in psychological distress in England, measured using the Slope Index of Inequality (SII) and Relative Index of Inequality (RII), 1991-2019 (Source: Health Survey for England)**

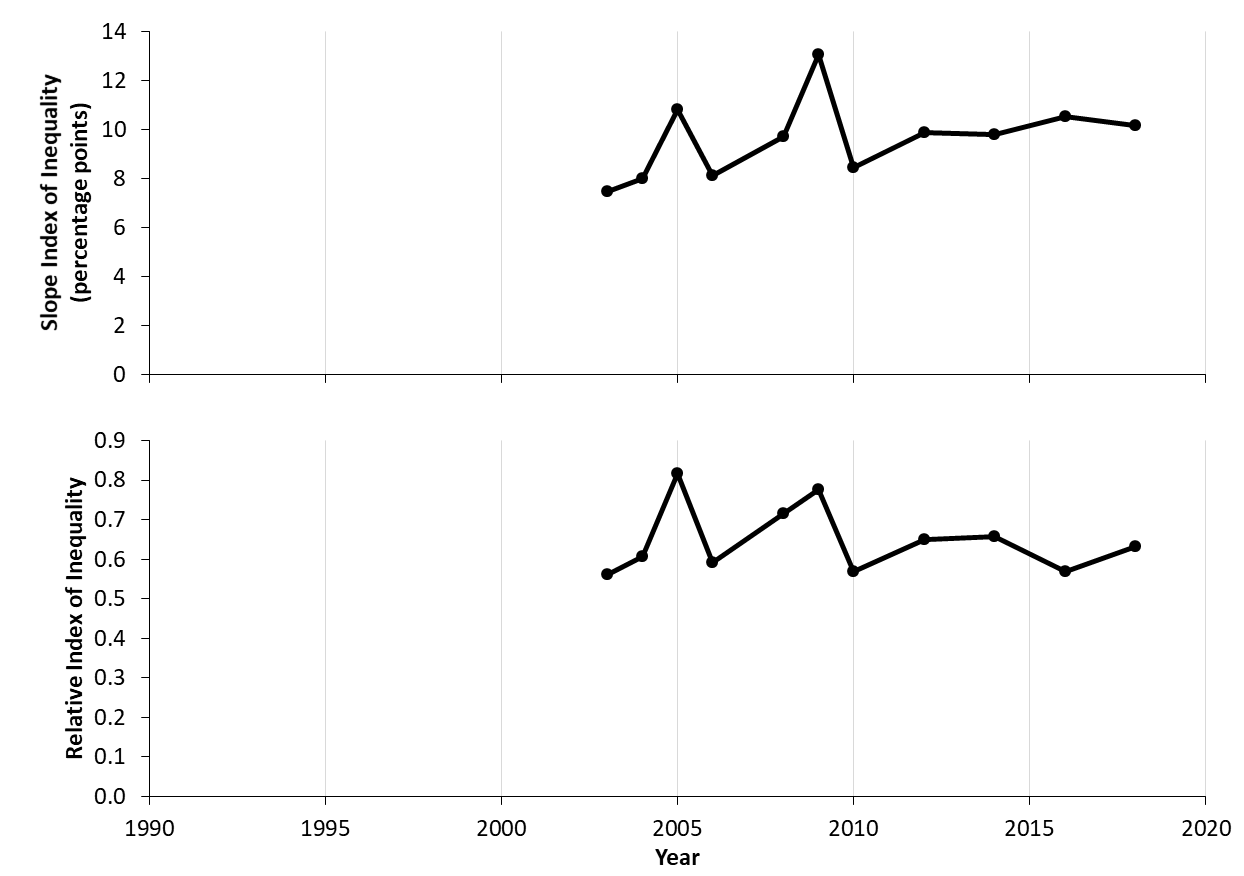

**Figure S2.6 – Trends in area deprivation-based inequalities in psychological distress in GB, males, 1991-2019 (Sources: Understanding Society, Scottish Health Survey, Health Survey for England)**
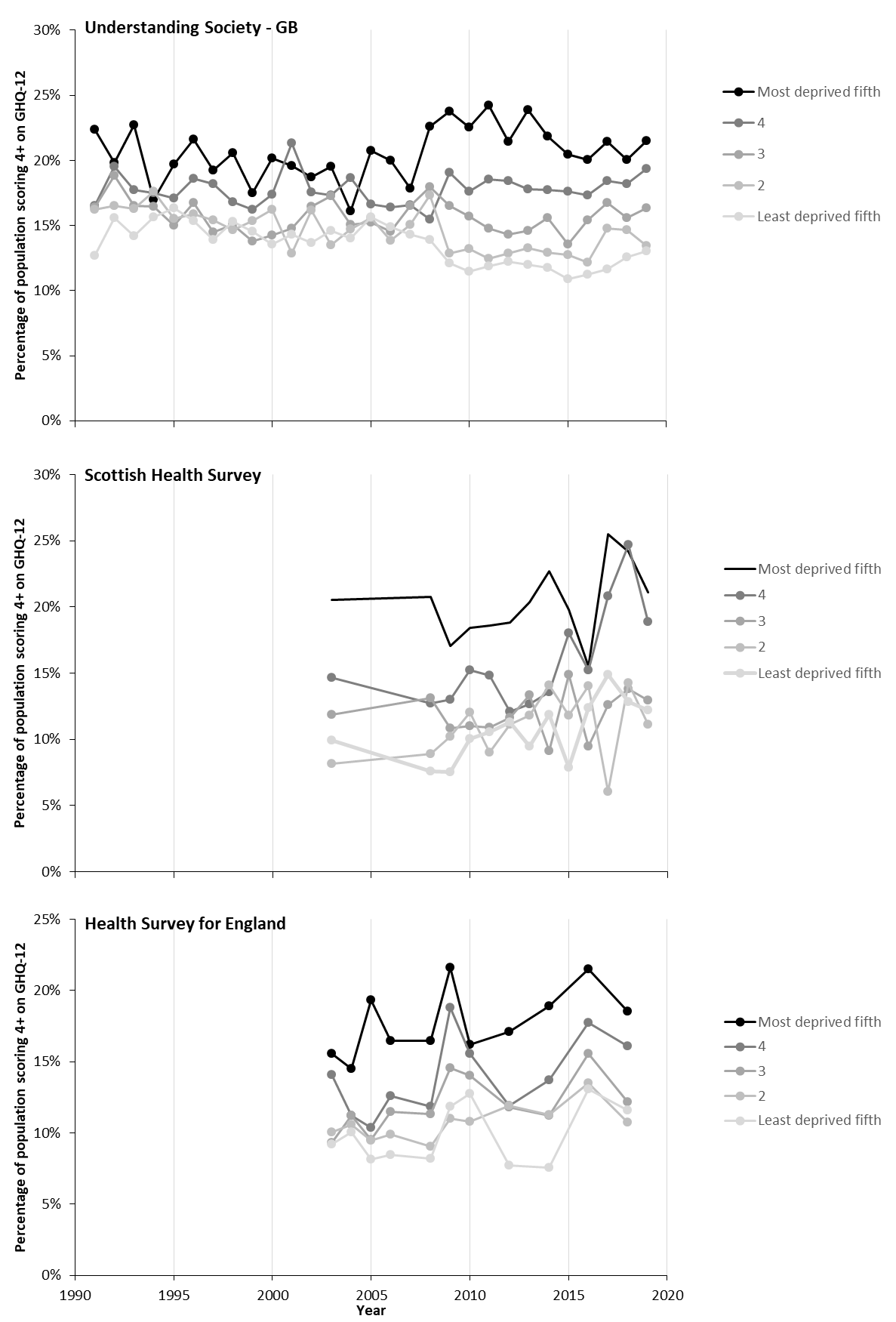

**Figure S2.7 – Trends in area deprivation-based inequalities in psychological distress in GB, females, 1991-2019 (Sources: Understanding Society, Scottish Health Survey, Health Survey for England)**
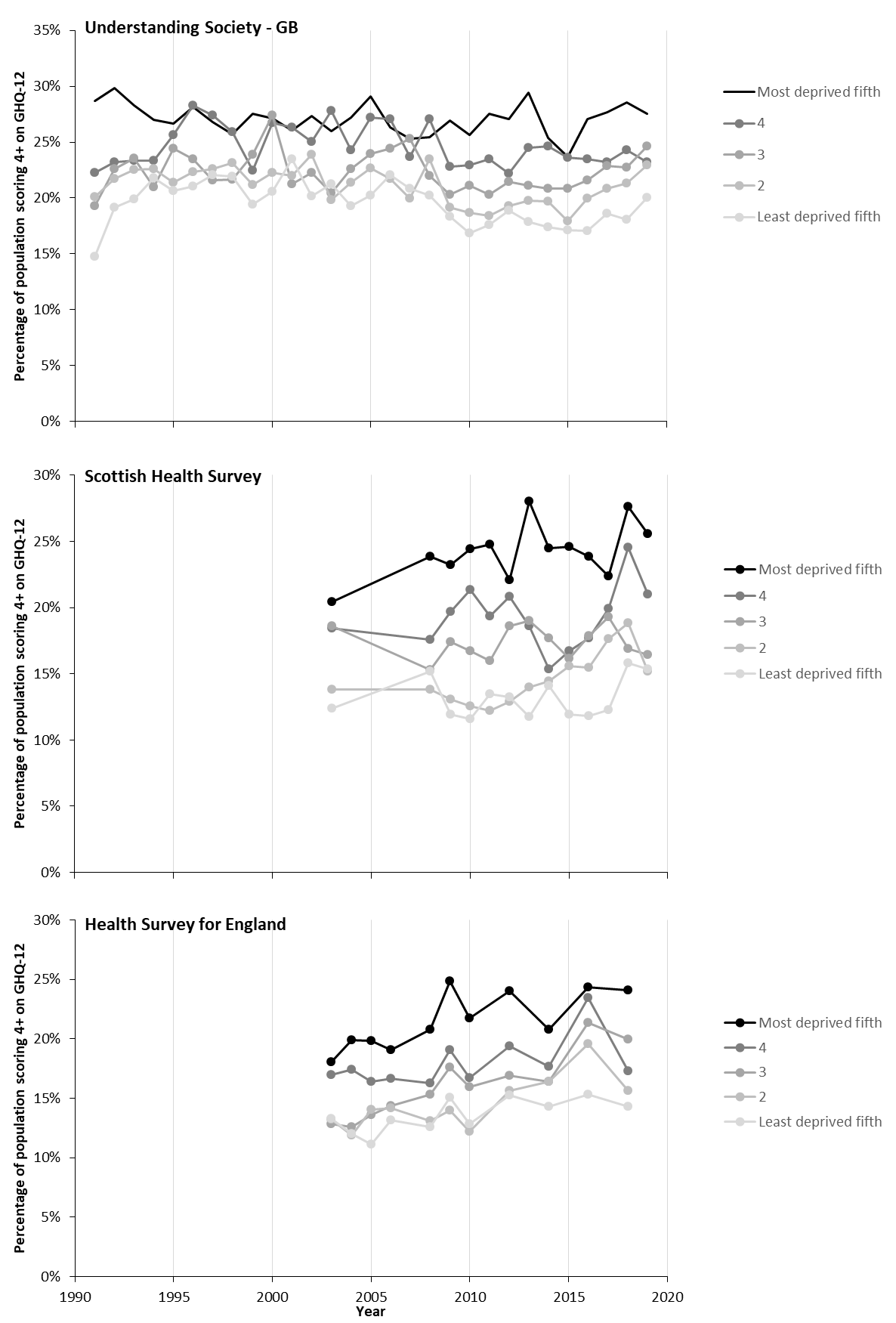

**Figure S2.8 – Trends in area deprivation-based inequalities in psychological distress in GB, males and females combined, 16-34 year olds, 1991-2019 (Sources: Understanding Society, Scottish Health Survey, Health Survey for England)**
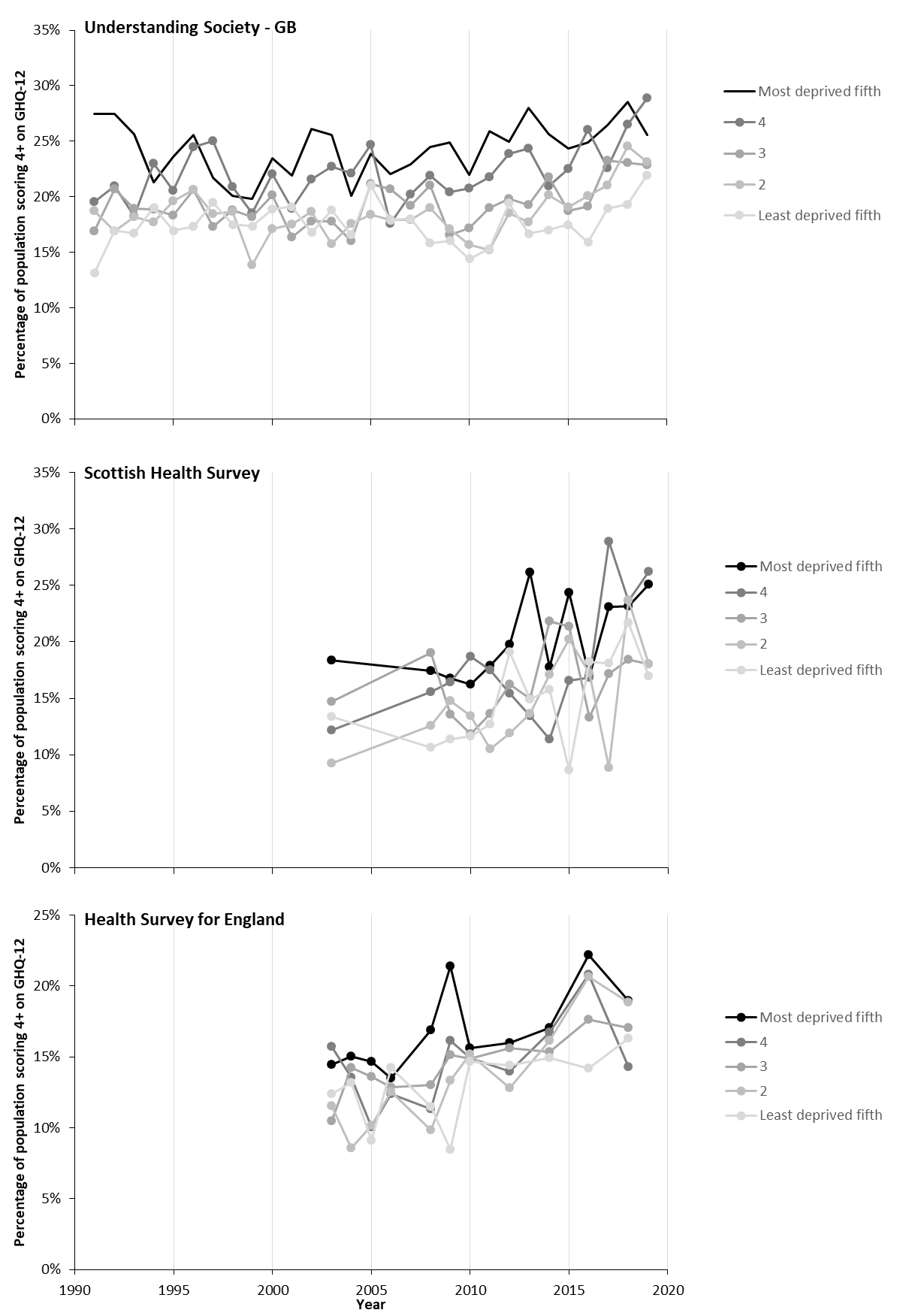

**Figure S2.9 – Trends in area deprivation-based inequalities in psychological distress in GB, males and females combined, 35-64 year olds, 1991-2019 (Sources: Understanding Society, Scottish Health Survey, Health Survey for England)**

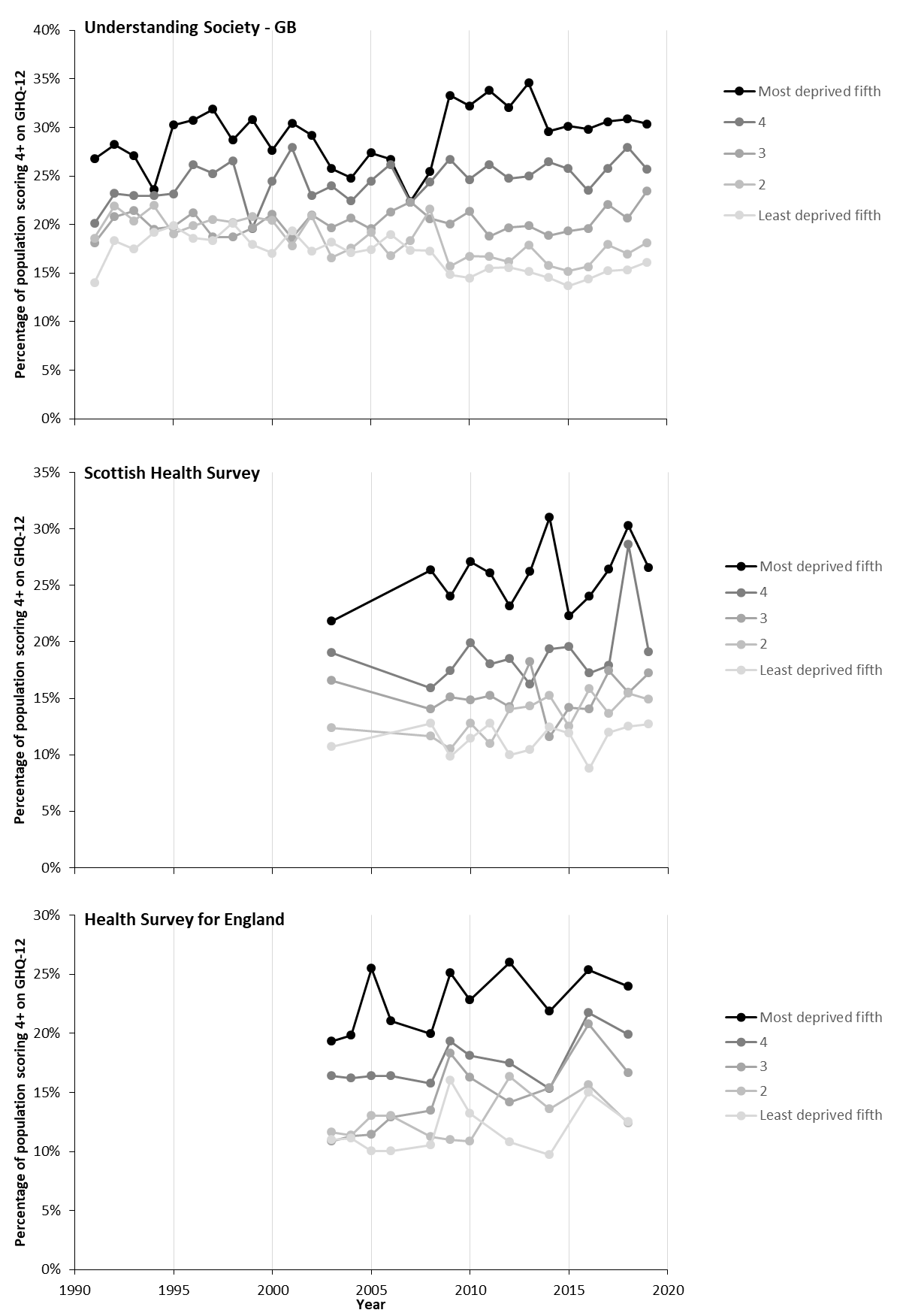

**Figure S2.10 – Trends in area deprivation-based inequalities in psychological distress in GB, males and females combined, 65+ year olds, 1991-2019 (Sources: Understanding Society, Scottish Health Survey, Health Survey for England)**
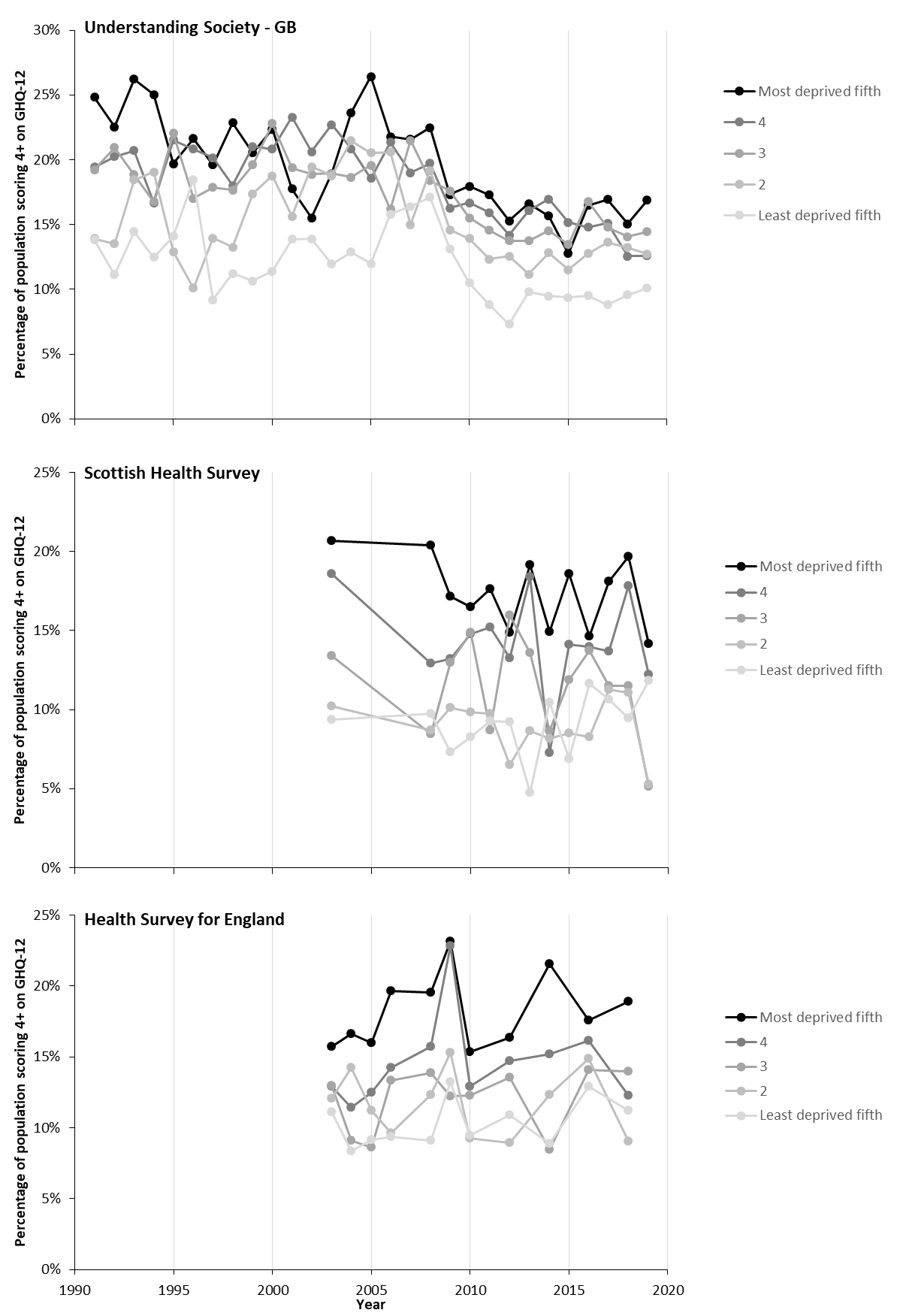

**Figure S2.11 – Trends and turning points (between 2010-2019) in the trends in prevalence of psychological distress for the total population, 1991-2019 (Sources: Understanding Society, Scottish Health Survey, Health Survey for England)**

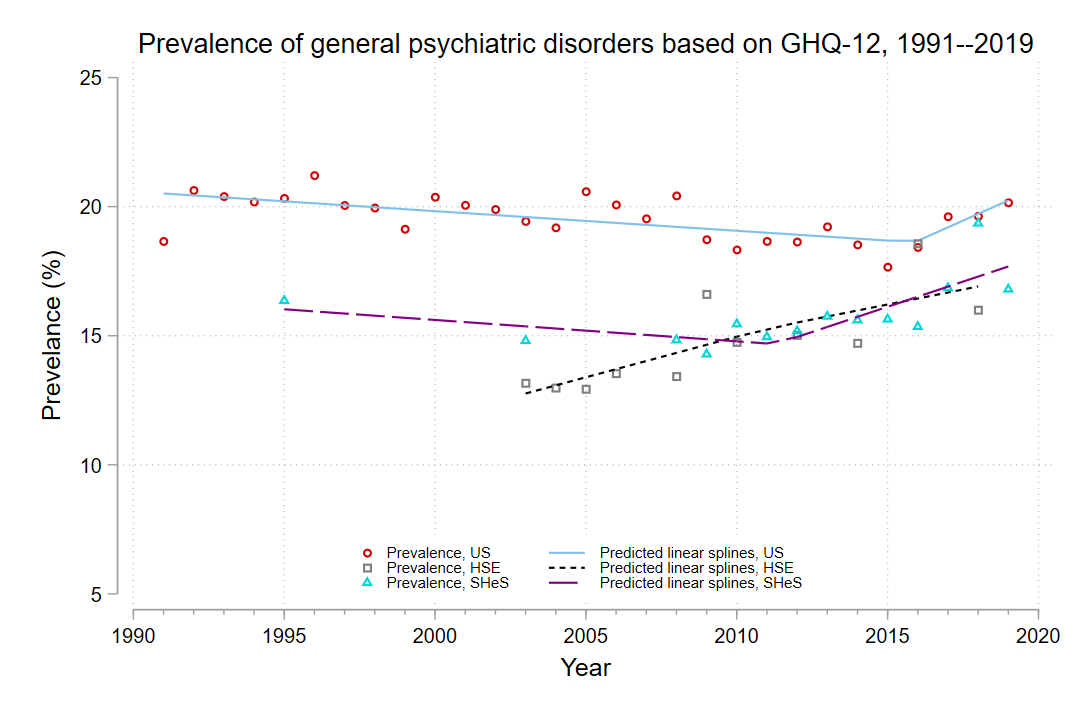

**Figure S2.12 – Trends and turning points (between 2010-2019) in the trends in prevalence of psychological distress by sex, 1991-2019 (Sources: Understanding Society, Scottish Health Survey, Health Survey for England)**
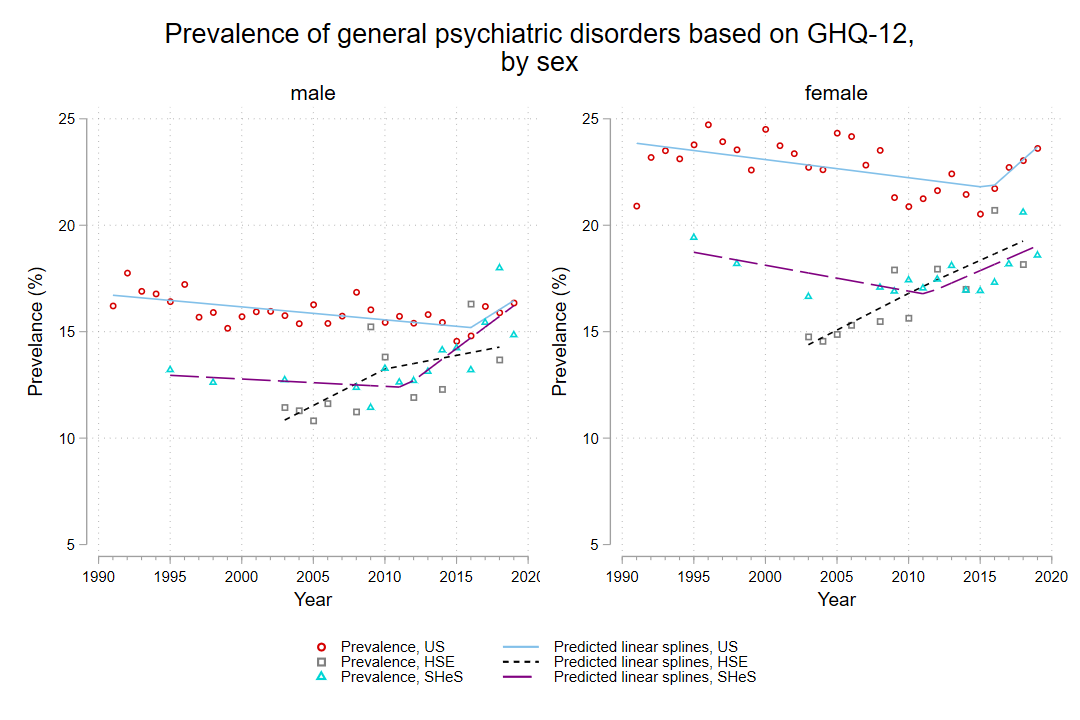

**Figure S2.13 – Trends and turning points (between 2010-2019) in the trends in prevalence of psychological distress by age strata, 1991-2019 (Sources: Understanding Society, Scottish Health Survey, Health Survey for England**

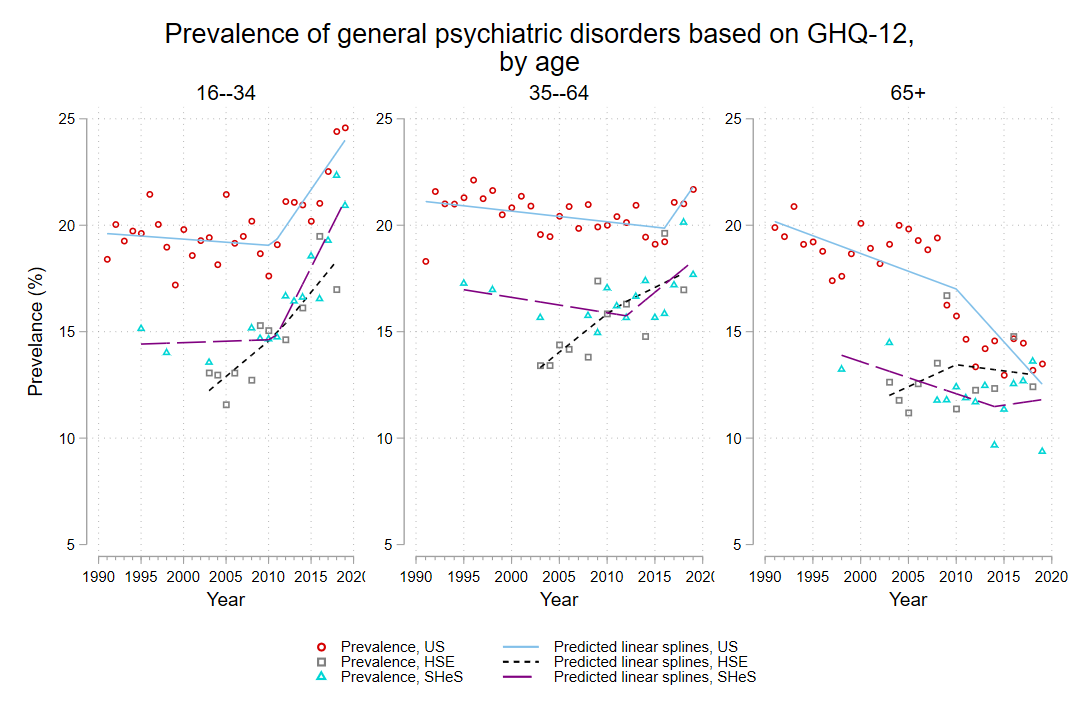

**Figure S2.14 – Trends and turning points (between 2010-2019) in the trends in prevalence of psychological distress by deprivation strata, 1991-2019 (Sources: Understanding Society, Scottish Health Survey, Health Survey for England**

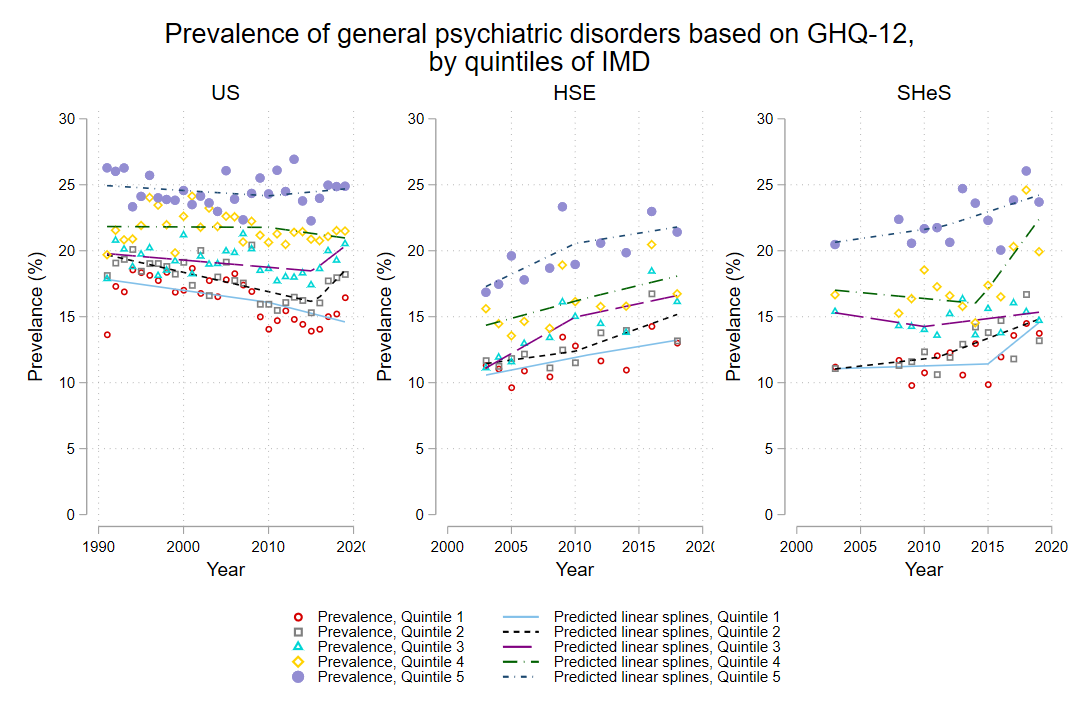
